# The additive effect of genetic modifiers on ALS prognosis: a population-based study

**DOI:** 10.1101/2022.09.25.22280338

**Authors:** Adriano Chiò, Cristina Moglia, Antonio Canosa, Umberto Manera, Maurizio Grassano, Rosario Vasta, Francesca Palumbo, Salvatore Gallone, Maura Brunetti, Marco Barberis, Fabiola De Marchi, Clifton Dalgard, Ruth Chia, Gabriele Mora, Barbara Iazzolino, Laura Peotta, Bryan Traynor, Lucia Corrado, Sandra D’Alfonso, Letizia Mazzini, Andrea Calvo

**Author notes:** Correspondence to: Adriano Chiò, “Rita Levi Montalcini” Department of Neuroscience, ALS Centre, University of Turin, Turin, 10126, Italy. These authors have equally contributed to this paper (Adriano Chiò, Cristina Moglia, Antonio Canosa, Sandra D’Alfonso, Letizia Mazzini, Andrea Calvo).

## Abstract

**Objective:** To determine if the co-presence of genetic polymorphisms related to ALS has additive effects on the course of the disease in a population-based cohort of Italian patients.

**Methods:** The study population includes 1245 ALS patients identified through the Piemonte Register for ALS, diagnosed between 2007 and 2016 and not carrying *SOD1, TARDBP* and *FUS* mutations. Controls were 766 age, sex, and geographically matched Italian subjects. We considered *UNC13A (*rs12608932), *CAMTA1 (*rs2412208), *SLC112A (*rs407135) and *ZNF512B (ZNF512B)* polymorphisms, as well as *ATXN2* polyQ intermediate repeats and *C9ORF72* GGGGCC intronic expansion.

**Results:** The variants in *C9orf72* (p=0.016), *ATXN2* (p<0.001) and *UNC13A* (p<0.001) were significantly related to survival in univariate analysis, while the other considered variants did not influence ALS outcome. However, in the Cox multivariable analysis, also *CAMTA1* emerged to be independently related to survival. When assessing the interaction by pairs of genes, we found that the presence of both detrimental alleles/expansion was correlated with significantly shorter survival compared to subjects non-carrying both detrimental alleles/expansions. Each association of pairs of detrimental alleles was characterized by specific clinical phenotypes.

**Conclusions:** we demonstrated that gene polymorphisms acting as genetic modifiers of ALS survival can act on their own or in unison. Overall, 54% of patients carried at least one detrimental common polymorphism or repeat expansion, highlighting the clinical impact of our findings. In addition, the identification of the synergic effects of modifier genes represents an essential clue for explaining ALS clinical heterogeneity and should be considered in designing and interpreting clinical trials.

**Key messages:** *What is already known on this topic:* Besides the disease-causing genes, several other genes have been reported to act as modifiers of ALS phenotype, especially patients’ survival. However, the interactions of these genes at clinical level have never been explored.

*What this study adds:* We demonstrated that gene polymorphisms and expansions acting as genetic modifiers of ALS survival can act on their own or in unison. Overall, 54% of patients carried at least one detrimental allele at common polymorphism or repeat expansion, highlighting the clinical impact of our findings.

*How this study might affect research, practice, or polic:* The identification of the synergic effects of modifier genes represents an essential clue for explaining ALS clinical heterogeneity, will have deep effects on clinical trial design and interpretation and support the inclusion of these polymorphisms in ALS genetic panels.

## Introduction

Amyotrophic lateral sclerosis (ALS) is a progressive degenerative disorder of the CNS, characterized by the involvement of upper motor neurons and lower motor neurons, as well as the cortical neurons of the frontotemporal cortices. ALS is considered a multifactorial disorder caused by an interaction between genetics and the environment.^1^ While relatively little is known about the environmental contributions to ALS, mutations in more than thirty genes have been linked to the disease, the most common being *C9orf72, SOD1, TARDBP* and *FUS*. Overall, the genetic etiology is known for about 70% of ALS patients with a familial history of ALS (fALS) and 10% of apparently sporadic ALS patients (sALS).^2^

Besides the disease-causing genes, several other genes have been reported to act as modifiers of ALS phenotype, especially patients’ survival. Among these, the most relevant are *UNC13A* (rs12608932 polymorphism),^3,4^ *CAMTA1* (rs2412208 polymorphism)^5^ *ATXN2* (intermediate polyQ repeats),^6,7^ *SLC11A2* (rs407135 polymorphism),^8^ and *ZNF512B* (rs2275294 polymorphism).^9^ Interestingly, *UNC13A* polymorphism has been demonstrated to be also a modifier of the response to drugs.^10^ These observations are important from the clinical trial perspective. Not only does it provide additional new targets for drug development, but it also suggests that these data should be incorporated into the clinical trial design; their effect on survival often equals the anticipated therapeutic effect, meaning balancing of genotypes in the treatment and placebo arms is needed to avoid false positive findings.

Despite these advances, it is not clear whether the various ALS genes interact in a manner that increases the risk of developing the disease. Therefore, this study aimed to determine if the co-presence of polymorphisms related to disease has additive effects on the course of ALS in a population-based cohort.

## Methods

The study population includes the 1,319 ALS patients identified through the Piemonte and Valle d’Aosta Register for ALS (PARALS), a prospective population-based register active in Northern Italy since 1995, diagnosed between 2007 and 2016, who underwent whole-genome sequencing on an Illumina HiSeqX10 sequencer. A total of 766 geographically-, age- and sex-matched Italian subjects were also sequenced. The characteristics of the epidemiological register have been reported elsewhere.^11^ Patients carrying mutations of *SOD1, TARDBP* and *FUS* genes (n=74) were excluded due to the heterogeneous clinical course of the different missense and nonsense mutations of these genes.^12^

### Whole Genome Analysis

Whole genome sequencing methods of 1029 ALS subjects and 766 controls have already been reported.^13^ An additional 290 ALS cases underwent whole genome sequencing as described elsewhere.^14^

### ATXN2 CAG and C9orf72 repeat analysis

*ATXN2* polyQ repeat in exon 1 (NM_002973.3) was amplified using a fluorescent primer and sized by capillary electrophoresis on an ABI3130 genetic analyzer (Applied Biosystems, Foster City, CA, USA).^6^ The presence of the *C9ORF72* intronic expansion was determined using an established repeat-primed PCR method described elsewhere.^15^

### Survival modifiers genes

For the present study, we considered the following genes reported to be related to ALS outcome: *UNC13A, CAMTA1, SLC11A2*, and *ZNF512B*. We also considered the interaction between these genes and *C9orf72* repeat expansion and *ATXN2* polyQ intermediate repeats, which also affects the survival of patients with ALS. Gene polymorphisms were dichotomized as follows: *UNC13A*^C/C^ vs *UNC13A*^A/A+A/C^; *CAMTA1*^G/G+G/T^ vs *CAMTA1*^T/T^; *SLC11A2*^A/C+C/C^ vs *SLC11A2*^A/A^; *ZNF512B*^C/C+C/T^ vs. *ZNF512B*^T/T^. The first allele(s) reported here is the detrimental one. All dichotomies were based on the original papers reporting the gene in ALS or subsequent studies^3,4,5,8,9^ and were confirmed in our cohort (data not shown).

### Clinical variables

ALSFRS-R mean monthly decline (ΔALSFRS-R) was calculated using the following formula: (48 – *ALSFRS-R score at diagnosis*)/(*months from onset to diagnosis*). Similarly, weight mean monthly decline (ΔWeight) was calculated as (*Weight at diagnosis* – *healthy body weight*)/*months from onset to diagnosis*. Finally, to have a proxy of disease spread, we calculated the mean monthly decline of King’s staging (ΔKing’s) as *(King’s staging at diagnosis)/(months from onset to diagnosis)*.

A total of 909 patients underwent an extensive cognitive battery at the time of diagnosis. These cases were classified into five categories according to the Consensus Criteria for diagnosing frontotemporal cognitive and behavioral syndromes in ALS.^16^ The battery assessed executive function, memory, visuospatial function, language, and social cognition, as well as anxiety and depression, and is reported in detail elsewhere.^17^

### Statistical analysis

First, the effect of survival of each gene was evaluated in isolation. Second, all genes were assessed together in Cox multivariable analysis. Third, the interaction of alleles on survival and other phenotypic characteristics was evaluated by pair of genes. Differences between continuous variables were assessed with the Mann-Whitney U test. Differences between discrete variables were assessed with the χ^2^ test. Survival was calculated with Kaplan-Meier curves and compared using the log-rank test, setting the onset date as day 0 and the date of death or tracheostomy as the primary endpoint. The last day of follow-up for censored cases was December 31, 2021.

Multivariable analysis for survival was performed with the Cox proportional hazards model (stepwise backward) with a retention criterion of a p-value less than 0.1. A p-value <0.05 was considered significant in the final model. The following variables were included in the model: age at onset (continuous), diagnostic delay (continuous), genetic gender (male versus female), site of onset (bulbar versus spinal), King’s staging, ΔALSFRS-R (continuous), FVC% at diagnosis (continuous), ΔWeight (continuous), ΔKing’s (continuous), and chronic obstructive pulmonary disease (COPD) (yes versus no). Analyses were conducted using the SPSS 28.0 statistical package (SPSS, Chicago, IL, USA).

### Ethical considerations

The study was approved by the Ethics Committees of the ALS Expert Centers of Torino and Novara (Comitato Etico Azienda Ospedaliero-Universitaria Città della Salute e della Scienza, Torino, and Comitato Etico Azienda Ospedaliero-Universitaria Maggiore della Carità, Novara #0038876). Patients and controls provided written informed consent before enrollment. The databases were anonymized according to Italian law for the protection of privacy.

### Data Availability Statement

Data will be available upon motivated request by interested researchers.

## Results

The study population included 1,245 ALS patients (n=689 males [55.3%], median age at onset = 67.8 years, IQR 59.9-74.4). The control participants were 766 age-, sex-, and geographically-matched Italian subjects, identified through patients’ general practitioners.

The frequency of the alleles of the examined polymorphisms is reported in Table 1. For *UNC13A*, the allele frequency was significantly different among cases and controls (p=0.037); allele frequency did not deviate from the Hardy-Weinberg Equilibrium among controls (p=0.29), while the deviation observed among patients (p=0.015) reflects the increased risk associated with the C allele. Also, for *ZNF512B*, allele frequency was significantly different among cases and controls (p=0.027); however, for patients and controls, the allele frequency of *ZNF512B* did not deviate from the Hardy-Weinberg Equilibrium. *CAMTA1* and *SLC11A2* allele frequencies were not significantly different among cases and controls (p=0.60 and p=0.33, respectively), and allele frequency did not deviate from the Hardy-Weinberg Equilibrium both in cases and controls. A total of 40 patients (3.2%) had *ATXN2* polyQ repeats ≥31, and 91 patients (7.3%) carried the *C9orf72* repeat expansion. The frequency of the combination of genetic polymorphisms and expansions did not deviate from the expected figures.

**Table 1.**
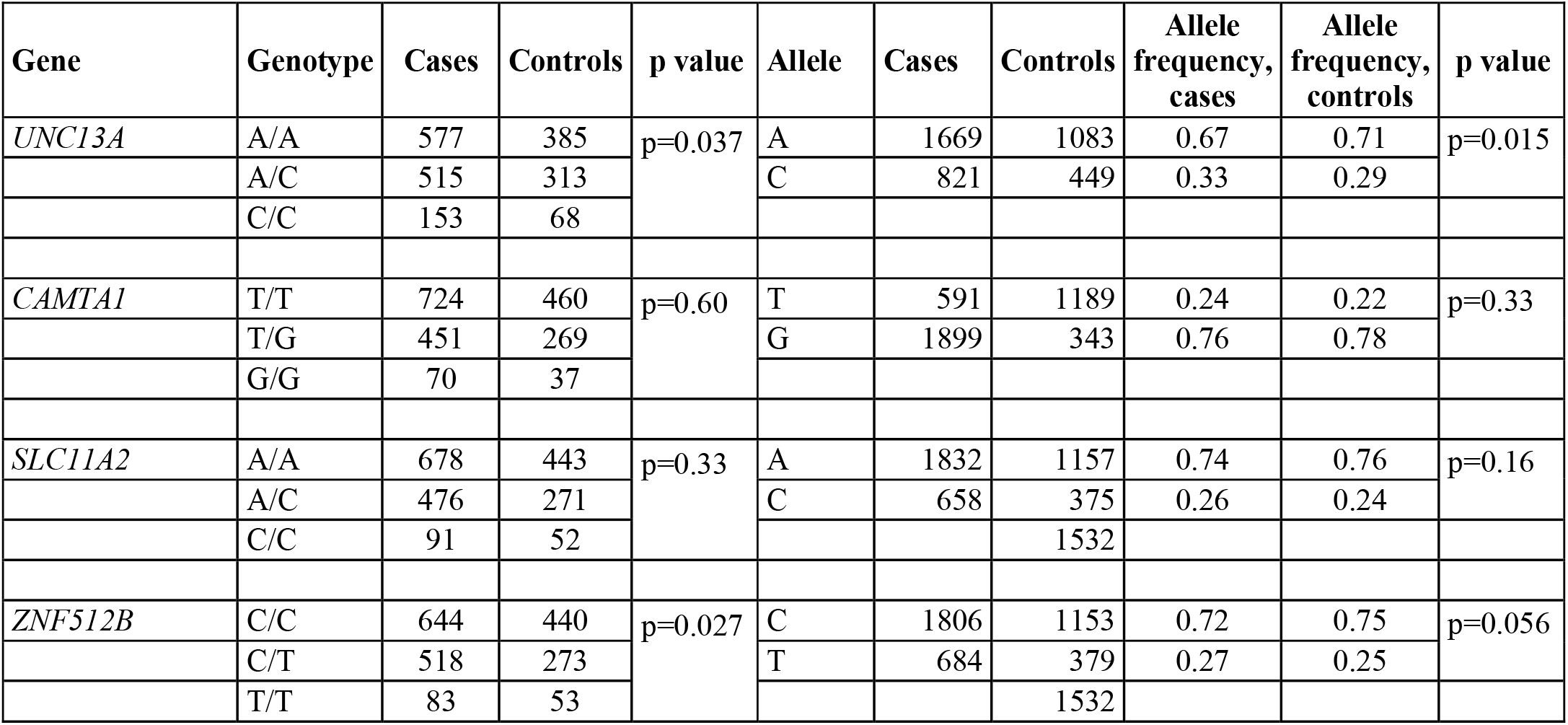
Frequency of genotypes and alleles of the studied polymorphisms.

| Gene | Genotype | Cases | Controls | p value | Allele | Cases | Controls | Allele frequency, cases | Allele frequency, controls | p value |
| --- | --- | --- | --- | --- | --- | --- | --- | --- | --- | --- |
| *UNC13A* | A/A | 577 | 385 | p=0.037 | A | 1669 | 1083 | 0.67 | 0.71 | p=0.015 |
|  | A/C | 515 | 313 |  | C | 821 | 449 | 0.33 | 0.29 |  |
|  | C/C | 153 | 68 |  |  |  |  |  |  |  |
| *CAMTA1* | T/T | 724 | 460 | p=0.60 | T | 591 | 1189 | 0.24 | 0.22 | p=0.33 |
|  | T/G | 451 | 269 |  | G | 1899 | 343 | 0.76 | 0.78 |  |
|  | G/G | 70 | 37 |  |  |  |  |  |  |  |
| *SLC11A2* | A/A | 678 | 443 | p=0.33 | A | 1832 | 1157 | 0.74 | 0.76 | p=0.16 |
|  | A/C | 476 | 271 |  | C | 658 | 375 | 0.26 | 0.24 |  |
|  | C/C | 91 | 52 |  |  |  | 1532 |  |  |  |
| *ZNF512B* | C/C | 644 | 440 | p=0.027 | C | 1806 | 1153 | 0.72 | 0.75 | p=0.056 |
|  | C/T | 518 | 273 |  | T | 684 | 379 | 0.27 | 0.25 |  |
|  | T/T | 83 | 53 |  |  |  | 1532 |  |  |  |

The examined variants in *C9orf72* (p=0.016), *ATXN2* (p<0.001) and *UNC13A* (p<0.001) were significantly related to survival in univariate analysis, while the variants in *CAMTA1* (p=0.231), *SLC11A2* (p=0.665) and *ZNF512B* (p=0.325) did not influence ALS outcome (Supplementary Figures 1 to 6). In the Cox multivariable analysis, *C9orf72* (Hazard Ratio [HR] 1.65, 95% c.i. 1.30-2.08, p<0.001), *ATXN2* (HR 1.65, 95% c.i. 1.18-2.29, p=0.003), *UNC13A* (HR 1.31, 95% c.i. 1.09-1.58, p=0.005), and *CAMTA1* (HR 1.13, 95% c.i. 1.001-1.30, p=0.048) were independently related to survival. Therefore, we assessed the combined effects of *C9orf72, ATXN2, UNC13A*, and *CAMTA1* on ALS outcome in patients with deleterious alleles or expansions compared to those without, on a pairwise basis.

When assessing the interaction by pairs of genes, we found that, in most gene pairs, the presence of both detrimental alleles/repeat expansion was correlated with significantly shorter survival compared to other cases. The partial exception was the interaction between *C9orf72* and *CAMTA1*, which was only marginally significant (p=0.052). Specifically, a total of 68 cases (5.5%) carried the *CAMTA1*^G/G+G/T^ and *UNC13A*^C/C^ alleles. Their median survival was 1.67 (1.16-3.08) years compared to 2.75 (1.67-5.26) for patients who did not carry detrimental alleles at both genes (p<0.001) (Figure 1). From the phenotypic perspective, patients with both *CAMTA1*^G/G+G/T^ and *UNC13A*^C/C^ alleles were characterized by a 4-year older age at onset, a more frequent bulbar onset, and a higher ΔWeight (Supplementary Table 1).

**Figure 1.**
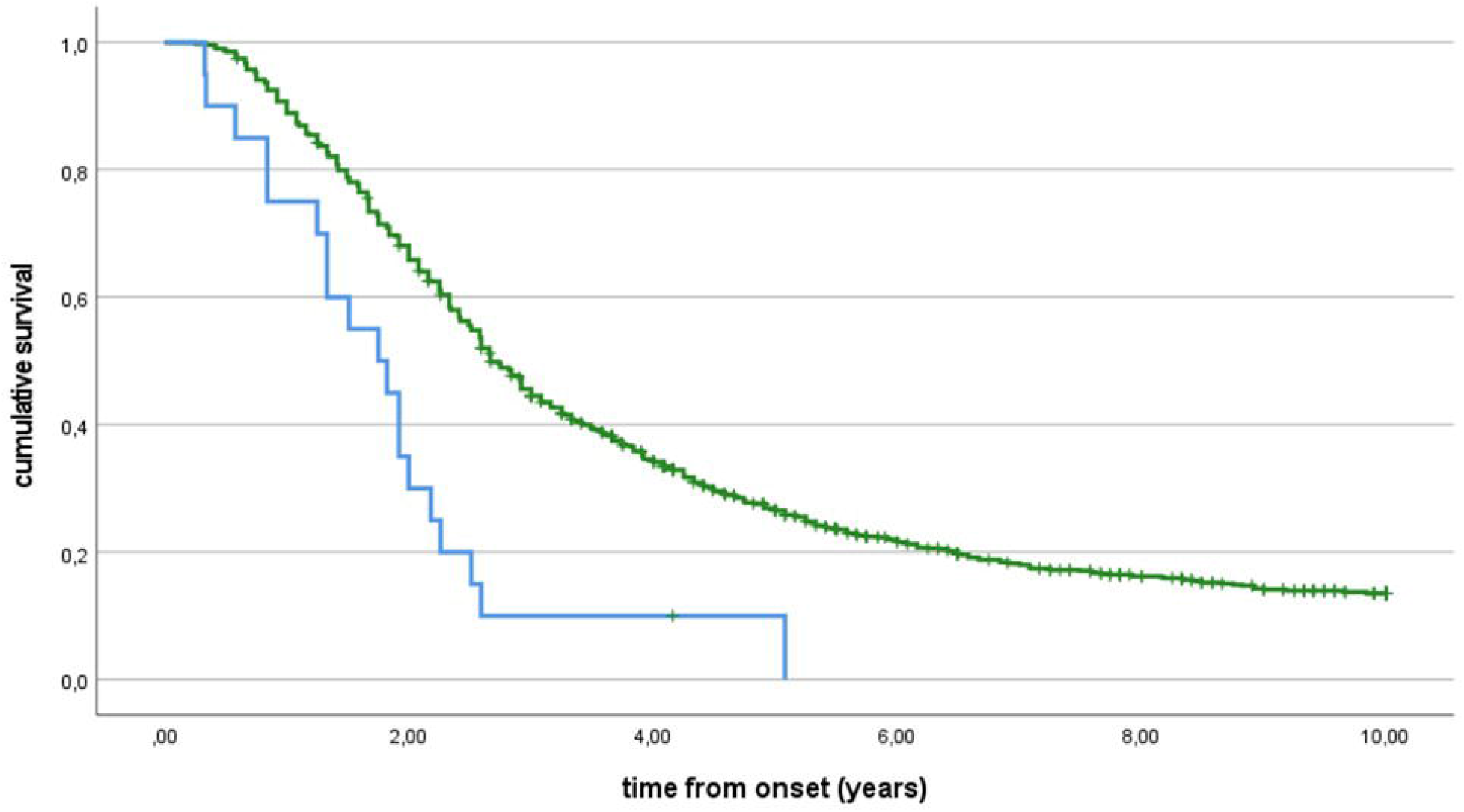
Survival curves (Kaplan Meier) for the interaction between *CAMTA1* rs2412208 polymorphism and UNC13A rs12608932 polymorphism. Median survival time: *CAMTA1*^G/G+G/T^ and *UNC13A*^C/C^ (68 cases, blue line) 1.67 years (1.16-3.08), *CAMTA1*^T/T^ and *UNC13A*^A/A+A/C^ (1177 cases, green line) 2.75 years (1.67-5.26), p<0.001. Ticks represent censored patients.

A total of 20 cases (1.6%) carried the *CAMTA1*^G/G+G/T^ alleles and the *ATXN2*^≥31^ intermediate polyQ repeats. Their median survival was 1.75 (0.84-2.18) years compared to 2.67 (1.67-5.25) for patients who did not carry detrimental alleles at both genes (p<0.001) (Figure 2). The phenotype of patients with both *CAMTA1*^G/G+G/T^ alleles and *ATXN2*^≥31^ CAG repeats was characterized by a more frequent bulbar onset and a higher ΔALSFRS-R and ΔKing’s (Supplementary Table 2).

**Table 2.** Patients’ survival. Cox multivariable analysis

| Factors | Values | Hazard Ratio (95% c.i.) | p value |
| --- | --- | --- | --- |
| Age at onset (years) | Per each year of age at onset | 1.029 (1.023-1.036) | <0.001 |
| Diagnostic delay | Per each month of delay | 0.955 (0.946-0.964) | <0.001 |
| Site of onset | Spinal | 1 [reference] | <0.001 |
|  | Bulbar | 1.484 (1.288-1.709) |  |
| ΔALSFRS-R | Per each point loss/month | 1.329 (1.267-1.394) | <0.001 |
| ΔKing's | Per each point loss/month | 1.663 (1.259-2.197) | <0.001 |
| *C9orf72* repeats | *C9ORF72*<sup>≤29</sup> | 1 [reference] | <0.001 |
|  | *C9ORF72*<sup>≥30</sup> | 1.645 (1.302-2.079) |  |
| *ATXN2* polyQ repeats | *ATXN2*<sup>≤30</sup> | 1 [reference] | 0.003 |
|  | *ATXN2*<sup>≥31</sup> | 1.645 (1.181-2.292) |  |
| *UNC13A* | *UNC13A*<sup>A/A+A/C</sup> | 1 [reference] | 0.005 |
|  | *UNC13A*<sup>C/C</sup> | 1.309 (1.087-1.578) |  |
| COPD | No | 1 [reference] | 0.011 |
|  | Yes | 1.330 (1.068-1.656) |  |
| *CAMTA1* | *CAMTA1*<sup>T/T</sup> | 1 [reference] | 0.048 |
|  | *CAMTA1*<sup>G/G+G/T</sup> | 1.135 (1.001-1.287) |  |

**Figure 2.**
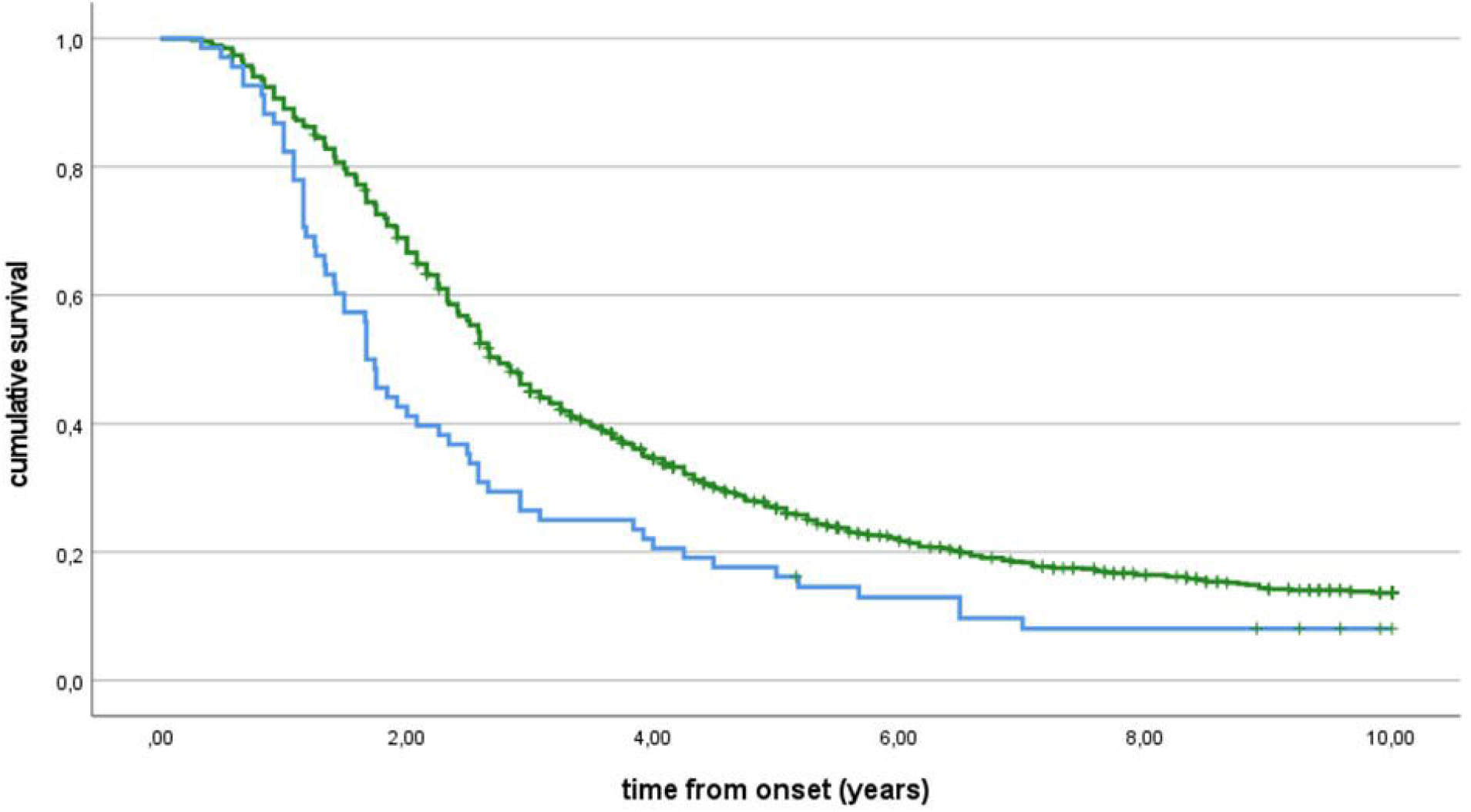
Survival curves (Kaplan Meier) for the interaction between *ATXN2* polyQ repeats and *CAMTA1* rs2412208 polymorphism. Median survival time: *ATXN2*^≥31^ and *CAMTA1*^G/G+G/T^ (20 cases, blue line) 1.75 years (0.84-2.18), *ATXN2*^≤30^ and *CAMTA1*^T/T^ (1225 cases, green line) 2.67 years (1.67-5.25), p<0.001. Ticks represent censored patients.

A total of 38 cases (3.1%) carried both the *CAMTA1*^G/G+G/T^ alleles and the *C9ORF72* repeat expansion. Their median survival was 2.33 (1.49-3.84) years compared to 2.67 (1.67-5.25) for patients who did not carry any of these detrimental alleles (p=0.052) (Figure 3). Patients with *CAMTA1*^G/G+G/T^ alleles and *C9ORF72*≥^30^ had an 8-year younger age at onset and were more frequently affected by co-morbid FTD (34.4% vs. 15.3%) (Supplementary Table 3).

**Figure 3.**
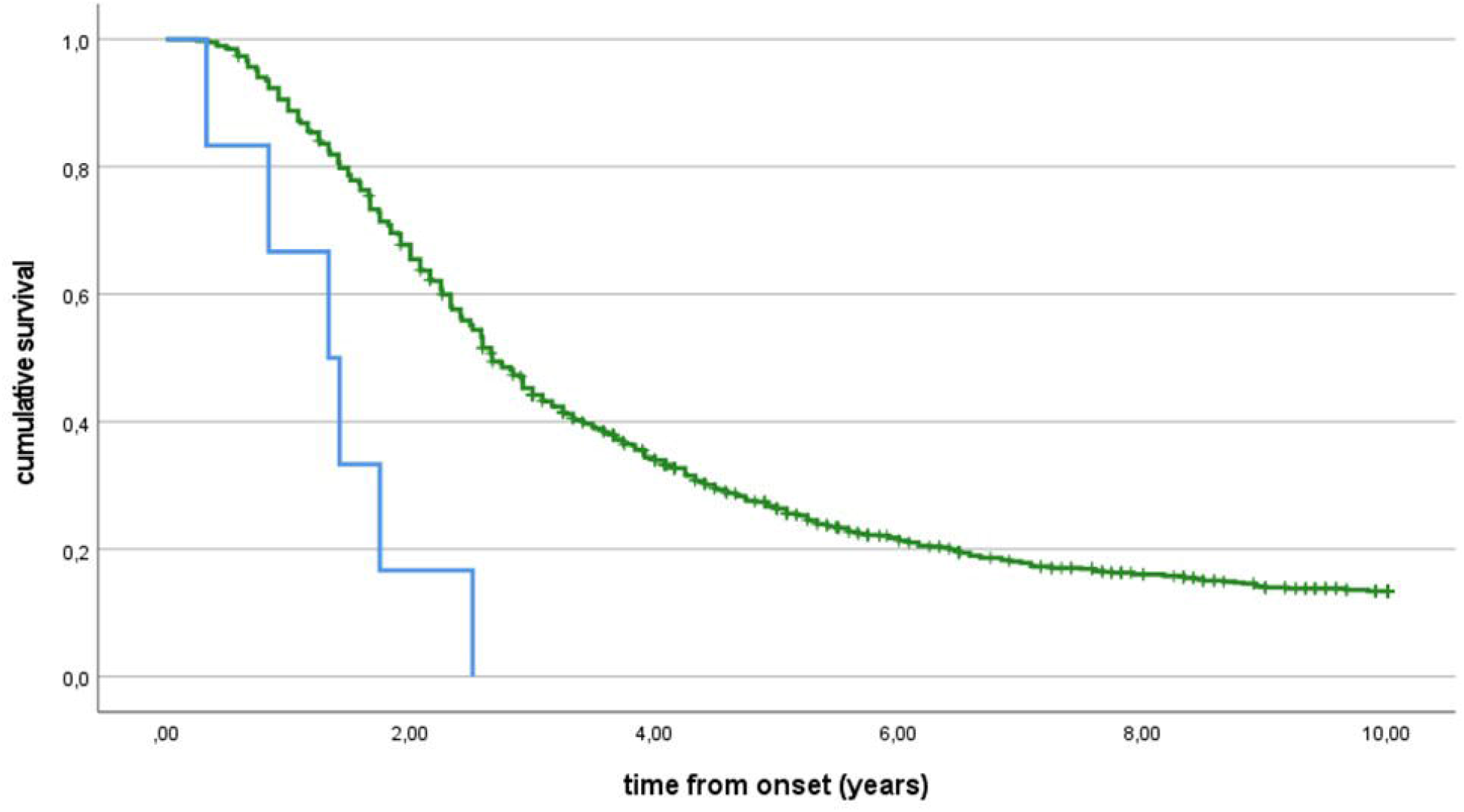
Survival curves (Kaplan Meier) for the interaction between *CAMTA1* rs2412208 polymorphism and *C9orf72* GGGGCC expansion. Median survival time: CAMTA1^G/G+G/T^ and *C9orf72*^≥30^ (38 cases, blue line) 2.33 years (1.49-3.84), CAMTA1^T/T^ and *C9orf72*^≤29^ (1207 cases, green line) 2.67 years (1.67-5.25), p=0.052. Ticks represent censored patients.

Six patients (0.5%) carried both *ATXN2*≥^31^ polyQ repeats and *UNC13A*^C/C^ alleles; their median survival time was 1.33 (0.84-1.75) vs. 2.67 (1.67-5.25) for those who carried *ATXN2*^≤30^ CAG repeats and *UNC13A*^A/A+A/G^ (p<0.001) (Figure 4).

**Figure 4.**
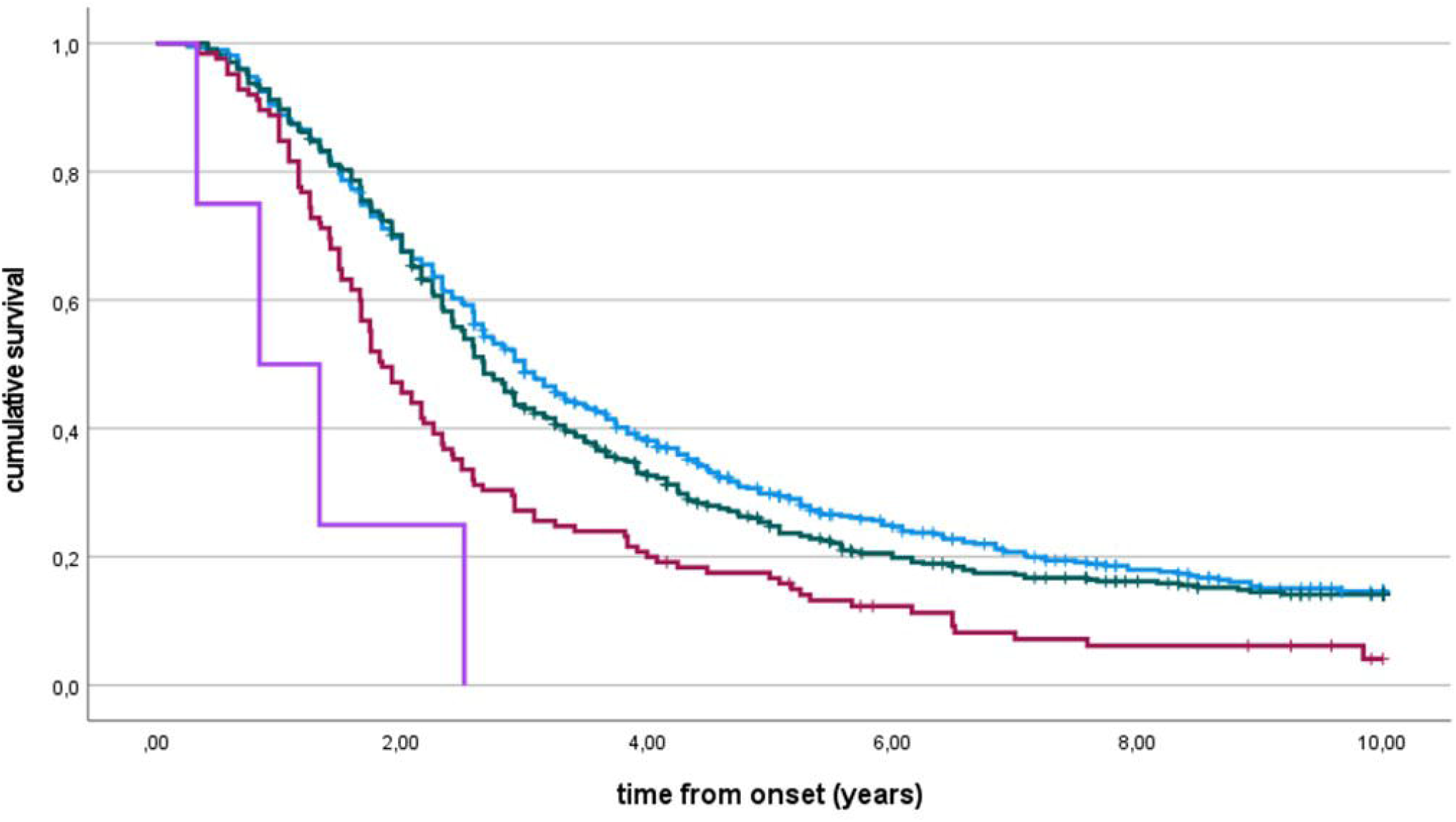
Survival curves (Kaplan Meier) for the interaction between *ATXN2* polyQ repeats and *UNC13A* rs12608932 polymorphism. Median survival time: *ATXN2*^≥31^ and *UNC13A*^C/C^ (6 cases, blue line) 1.33 years (0.84-1.75) *ATXN2*^≤30^ and *UNC13A*^A/A+A/C^ (1239 cases, green line) 2.67 years (1.67-5.25), p <0.001. Ticks represent censored patients.

Five patients (0.4%) carried both *C9ORF72*^≥30^ and *UNC13A*^C/C^ alleles; their median survival time was 1.66 (1.41-2.16) vs. 2.67 (1.67-5.25) for those who carried *C9ORF72*^≥30^ and *UNC13A*^A/A+A/G^ (p<0.019) (Figure 5).

**Figure 5.**
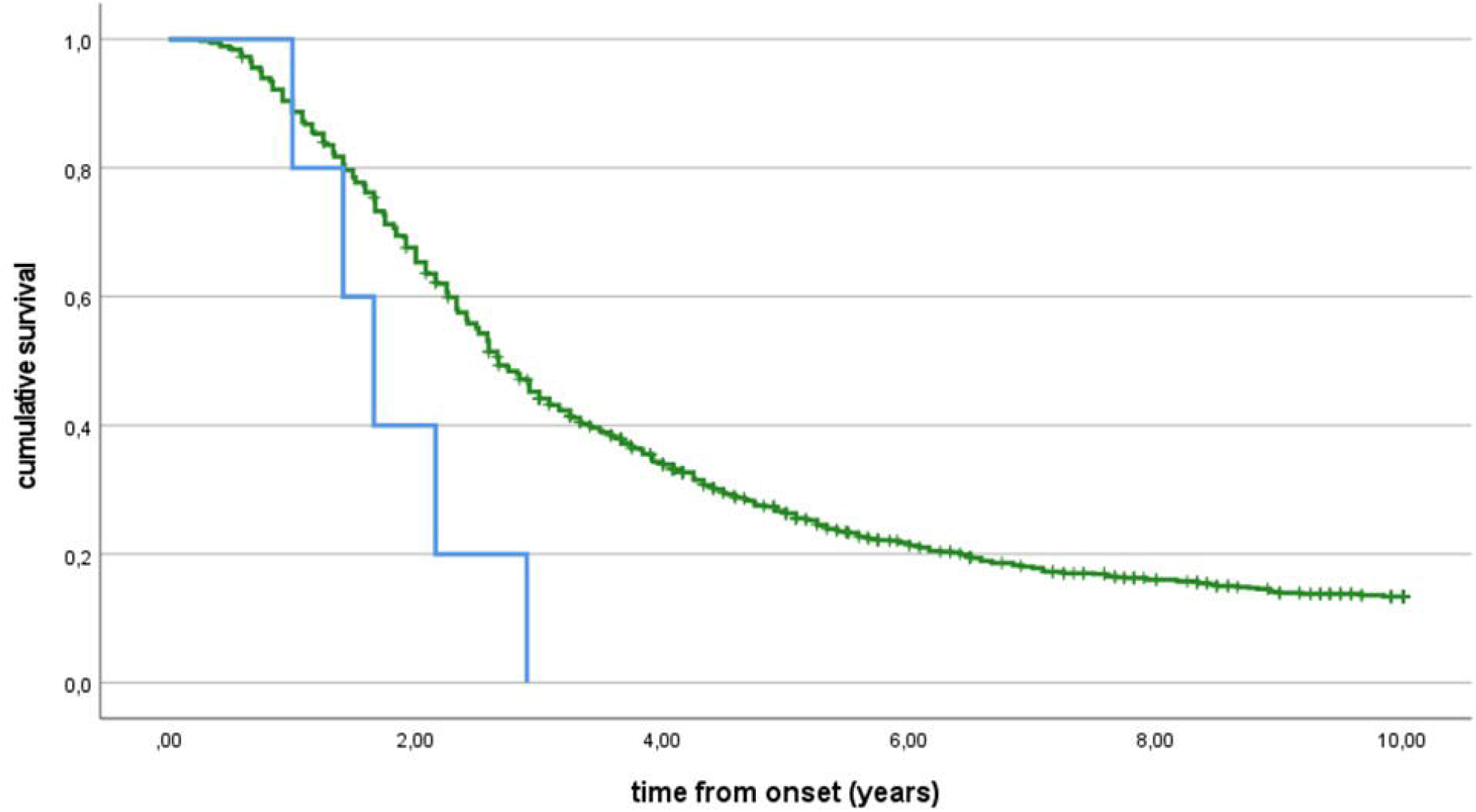
Survival curves (Kaplan Meier) for the interaction between *C9orf72* GGGCC expansion and UNC13A rs12608932 polymorphism. Median survival time: *C9orf72*^≥30^ and *UNC13A*^C/C^ (5 cases, blue line) 1.66 years (1.41-2.16), *C9orf72*^≤29^ and *UNC13A*^A/A+A/C^ (1240 cases, green line) 2.67 years (1.67-5.25), p<0.019. Ticks represent censored patients.

Finally, in the present cohort, no cases with *C9orf72* expansion carried also an *ATXN2* intermediate repeat expansion.

We also evaluated the effect on ALS outcome in patients with one, two or three deleterious alleles. The 573 patients (46.0%) with no deleterious allele had a median survival time of 3.0 years (1.67-5.92) compared to 2.67 years (1.75-5.0) for those carrying one deleterious polymorphisms/expansions (543 cases, 43.6%), 1.84 years (1.25-3.25) for those carrying two deleterious polymorphisms/expansions (125 cases, 10.0%), and 0.84 years (0.33-1.33) for the 4 (0.3%) patients carrying three polymorphisms (p<0.001). The corresponding survival curves are reported in Figure 6.

**Figure 6.** Survival curves (Kaplan Meier) according to the number of deleterious polymorphisms and or repeat expansions carries by each patient. Median survival time: patients with deleterious polymorphism (573, 46.0%, blue line) 3.0 years (1.67-5.92); patients with one deleterious polymorphism (543, 43.6%, green line), 2.67 years (1.75-5.0); patients carrying two deleterious polymorphisms (125 cases, 10.0%, red line) 1.84 years (1.25-3.25); patients carrying three deleterious polymorphisms (4, 0.3%, violet line) 0.84 years (0.33-1.33), p<0.001. Ticks represent censored patients.

## Discussion

In our population-based cohort, we have found that the co-presence of selected detrimental alleles at common polymorphisms or repeat expansions that individually are detrimental to survival in patients with ALS has an additive effect. In particular, the co-presence of *CAMTA1*^G/G+G/T^ polymorphisms with either *UNC13A*^C/C^ polymorphism or *ATXN2* polyQ intermediate expansion or *C9ORF72* expansion was related to a significantly worse patients’ outcome. This effect was also found when assessing the co-presence of *UNC13A*^C/C^ polymorphism with either *C9orf72* GGGGCC expansion or *ATXN2* polyQ intermediate expansion. In our cohort, 672 patients (54%) carried at least one deleterious polymorphism/expansion.

Genetic modifiers of ALS phenotype have been generally studied in isolation.^3,4,5,8,9^ Notable exceptions are two studies reporting that the co-occurrence of the *C9orf72* repeat expansion and *UNC13A*^C/C^ polymorphism significantly worsened the prognosis of patients with ALS.^18,19^ However, identifying the mechanisms underlying the wide phenotypic heterogeneity of ALS, which hinders the discovery of effective therapies,^2^ remains one of the significant unmet goals of ALS research. ALS heterogeneity is likely due to an interplay between genetics, age, gender,^1,20^ and environmental factors, both related to lifestyle (i.e., physical activity, smoking)^21,22^ and internal causes (i.e., lipid metabolism, gut microbiome).^23-25^ In this study, we have shown that another element determining ALS phenotypic heterogeneity is the co-presence of two or more different genetic modifiers of survival.

CAMTA1^G/G+G/T^ alleles in our cohort appear to interact with all other examined genes in shortening ALS survival. The interaction between *CAMTA1*^G/G+G/T^ and *UNC13A*^C/C^ polymorphisms, accounting for 68 patients (5.5%), besides the strong negative effect on survival, is also phenotypically characterized by an older age at onset, a more frequent bulbar onset, and a higher reduction of weight (ΔWeight). The interaction between *CAMTA1*^G/G+G/T^ and *ATXN2*^≥31^ (20 cases, 1.6%) is characterized by an increased ΔALSFRS-R and ΔKing’s, indicating a faster spreading and worsening of motor symptoms. Finally, patients with both *CAMTA1*^G/G+G/T^ and *C9ORF72*^≥30^ were younger and had an increased ΔKing’s. The number of cases with the interaction between *UNC13A*^C/C^ and *C9ORF72*^≥30^ (5 cases, 0.4%) and *UNC13A*^C/C^ and *ATXN2*^≥31^ (6 cases, 0.5%) was too low to detect any significant phenotypic difference.

The biological reasons for these interactions remain to be elucidated. The proteins encoded by these genes may interact at a molecular level. It has been reported that TDP-43 protein, cytoplasmic inclusions of which are a pathological hallmark of the disease, enhances translation of *CAMTA1* and *Mig12* via a gain-of-function mechanism operating through their 5′UTRs;^26^ however, this paper did not assess if the occurrence of *CAMTA1*^G/G+G/T^ polymorphism differentially influences the observed effect. More recently, it has been shown that TDP-43 represses a cryptic exon-splicing event in *UNC13A*, causing a reduction in *UNC13A* protein expression.^27,28^ In addition, the C/C genetic variation in *UNC13A* promotes cryptic exon inclusion upon nuclear depletion of TDP-43.^27,28^ Independently from the previously reported mechanism, *CAMTA1* was found to be a relevant ‘Master Regulator’ of neurodegenerative disease transcriptional programs in a cultured motor neuron-based ALS model.^29^

This study is not without limitations. First, not all patients were tested for cognitive function, limiting the possibility of assessing the genetic interactions on cognition. However, the clinical characteristics of tested and non-tested patients were similar, excluding a selection bias. Second, very few patients carried both *ATXN2*^≥31^ CAG repeats and *UNC13A*^C/C^ detrimental alleles or*C9ORF72*^≥30^ and *UNC13A*^C/C^ detrimental alleles, reducing the possibility of assessing their phenotype and limiting the power of these analyses. Similarly, due to the limited number of patients with more than two polymorphisms, we could not evaluate the effect of their association. Larger patient cohorts are necessary to analyze these combinations and to calculate a polygenic risk score for survival. A remarkable aspect of our study is its population-based nature since it included some 75% of incident patients in the Piemonte region.^13^ It has been demonstrated that prevalent and incident populations strongly differ from the clinical point of view, including survival, supporting the notion that studies derived from epidemiological cohorts better represent the ALS population.^30,31^

We demonstrated that gene polymorphisms and expansions acting as genetic modifiers of ALS survival can act on their own or in unison. Overall, 54% of patients carried at least one detrimental allele at common polymorphism or repeat expansion, highlighting the clinical impact of our findings. This observation has several implications. First, identifying the synergic effects of modifier genes represents an essential clue for explaining ALS clinical heterogeneity. Second, the additive effect of these polymorphisms is likely to have profound effects on clinical trial design and interpretation, in particular for relatively common combinations, such as the association of *CAMTA1*^G/G+G/T^ and *UNC13A*^C/C^ polymorphisms which accounted in our series for 5.5% of patients and reduced patients’ survival by more than one year. Third, our study indicates that polymorphisms acting as phenotypic modifiers should be included in ALS genetic panels to provide patients and their families with a better prediction of the course of the disease and improve the planning of therapeutic interventions.

## Data Availability

Data will be available upon motivated request by interested researchers.

https://www.neuroscienze.unito.it

## Author Contributions

Dr Chiò had full access to all of the data in the study and takes responsibility for the integrity of the data and the accuracy of the data analysis.

### Study concept and design

Chiò, Moglia, Canosa, Manera, Grassano, Vasta, Palumbo, Gallone, Brunetti, Barberis, De Marchi, Dalgard, Chia, Mora, Traynor, Corrado, D’Alfonso, Mazzini, Calvo

### Acquisition, analysis, or interpretation of data

Chiò, Moglia, Canosa, Manera, Grassano, Vasta, Palumbo, Gallone, Brunetti, Barberis, De Marchi, Dalgard, Chia, Mora, Traynor, Corrado, D’Alfonso, Mazzini, Calvo

### Drafting of the manuscript

Chiò, Moglia, Canosa, Traynor, D’Alfonso, Mazzini, Calvo

### Critical revision of the manuscript for important intellectual content

Chiò, Moglia, Canosa, Manera, Grassano, Vasta, Palumbo, Gallone, Brunetti, Barberis, De Marchi, Dalgard, Chia, Mora, Traynor, Corrado, D’Alfonso, Mazzini, Calvo

### Obtained funding

Traynor, Chiò

### Administrative, technical, or material support

Grassano, Palumbo, Gallone, Brunetti

### Study supervision

Chiò, Moglia, Canosa, D’Alfonso, Mazzini, Calvo

### Funding

This work was supported by the Italian Ministry of Health (Ministero della Salute, Ricerca Sanitaria Finalizzata, grant RF-2016-02362405); the Progetti di Rilevante Interesse Nazionale program of the Ministry of Education, University and Research (grant 2017SNW5MB); the European Commission’s Health Seventh Framework Programme (FP7/2007–2013 under grant agreement 259867); the Horizon 2020 Programme (project Brainteaser under grant agreement 101017598); and the Joint Programme–Neurodegenerative Disease Research (Strength, ALS-Care and Brain-Mend projects), granted by Italian Ministry of Education, University and Research. This work was supported in part by the Intramural Research Programs of the National Institute on Aging (grant Z01-AG000949-02). This study was performed under the Department of Excellence grant of the Italian Ministry of University and Research to the “Rita Levi Montalcini” Department of Neuroscience, University of Torino, Italy, and to the Department of Health Sciences, University of Eastern Piedmont, Novara, Italy. The funders had no role in data collection or analysis and did not participate in writing or approving the manuscript.

## References

1. Vasta R, Chia R, Traynor BJ, Chiò A. Unraveling the complex interplay between genes, environment, and climate in ALS. EBioMedicine. 2022 Jan;75:103795.

2. Goutman SA, Hardiman O, Al-Chalabi A, et al. Recent advances in the diagnosis and prognosis of amyotrophic lateral sclerosis. Lancet Neurol. 2022 May;21(5):480–493

3. Diekstra FP, van Vught PW, van Rheenen W, et al. UNC13A is a modifier of survival in amyotrophic lateral sclerosis. Neurobiol Aging. 2012 Mar;33(3):630.e3-8.

4. Chiò A, Mora G, Restagno G et al. UNC13A influences survival in Italian amyotrophic lateral sclerosis patients: a population-based study. Neurobiol Aging. 2013 Jan;34(1):357.e1-5.

5. Fogh I, Lin K, Tiloca C, et al. Association of a Locus in the CAMTA1 Gene With Survival in Patients With Sporadic Amyotrophic Lateral Sclerosis. JAMA Neurol. 2016 Jul 1;73(7):812–20.

6. Chiò A, Calvo A, Moglia C, et al. ATXN2 polyQ intermediate repeats are a modifier of ALS survival. Neurology. 2015 Jan 20;84(3):251–8.

7. Chio A, Moglia C, Canosa A, et al. Exploring the phenotype of Italian patients with ALS with intermediate ATXN2 polyQ repeats. J Neurol Neurosurg Psychiatry. 2022 Aug 25:jnnp-2022-329376. doi: 10.1136/jnnp-2022-329376.

8. Blasco H, Vourc’h P, Nadjar Y, et al; French ALS study group. Association between divalent metal transport 1 encoding gene (SLC11A2) and disease duration in amyotrophic lateral sclerosis. J Neurol Sci. 2011 Apr 15;303(1-2):124–7.

9. Jiang H, Yang B, Wang F, et al. Association of Single Nucleotide Polymorphism at rs2275294 in the ZNF512B Gene with Prognosis in Amyotrophic Lateral Sclerosis. Neuromolecular Med. 2021 Jun;23(2):242–246.

10. van Eijk RPA, Jones AR, Sproviero W, et al. Meta-analysis of pharmacogenetic interactions in amyotrophic lateral sclerosis clinical trials. Neurology. 2017 Oct 31;89(18):1915–1922.

11. Chiò A, Mora G, Moglia C, et al. Secular Trends of Amyotrophic Lateral Sclerosis: The Piemonte and Valle d’Aosta Register. JAMA Neurol. 2017 Sep 1;74(9):1097–1104.

12. Berdyñski M, Miszta P, Safranow K, et al. SOD1 mutations associated with amyotrophic lateral sclerosis analysis of variant severity. Sci Rep. 2022 Jan 7;12(1):103.

13. Grassano M, Calvo A, Moglia C, et al. Mutational Analysis of Known ALS Genes in an Italian Population-Based Cohort. Neurology. 2021 Jan 26;96(4):e600–e609.

14. Chia R, Sabir MS, Bandres-Ciga S, et al. Genome sequencing analysis identifies new loci associated with Lewy body dementia and provides insights into its genetic architecture. Nat Genet. 2021 Mar;53(3):294–303. doi: 10.1038/s41588-021-00785-3.

15. Chiò A, Calvo A, Mazzini L, et al. Extensive genetics of ALS: a population-based study in Italy. Neurology 2012; 79:1983-9.

16. Strong MJ, Abrahams S, Goldstein LH, et al. Revised diagnostic criteria. Amyotroph Lateral Scler Frontotemporal Degener 2017;18:153–174.

17. Iazzolino B, Peotta L, Zucchetti JP, et al. Differential Neuropsychological Profile of Patients With Amyotrophic Lateral Sclerosis With and Without C9orf72 Mutation. Neurology. 2021 Jan 5;96(1):e141–e152..

18. van Blitterswijk M, Mullen B, Wojtas A, et al. Genetic modifiers in carriers of repeat expansions in the C9ORF72 gene. Mol Neurodegener. 2014 Sep 20;9:38.

19. Dekker AM, Seelen M, van Doormaal PT, et al. Large-scale screening in sporadic amyotrophic lateral sclerosis identifies genetic modifiers in C9orf72 repeat carriers. Neurobiol Aging. 2016 Mar;39:220.e9-15.

20. Chiò A, Moglia C, Canosa A, et al. ALS phenotype is influenced by age, sex, and genetics: A population-based study. Neurology. 2020 Feb 25;94(8):e802–e810.

21. Calvo A, Canosa A, Bertuzzo D, et al. Influence of cigarette smoking on ALS outcome: a population-based study. J Neurol Neurosurg Psychiatry. 2016 Nov;87(11):1229–1233.

22. Rosenbohm A, Peter R, Dorst J, et al. Life Course of Physical Activity and Risk and Prognosis of Amyotrophic Lateral Sclerosis in a German ALS Registry. Neurology. 2021 Nov 9;97(19):e1955–e1963.

23. Bandres-Ciga S, Noyce AJ, Hemani G, et al. Shared polygenic risk and causal inferences in amyotrophic lateral sclerosis. Ann Neurol. 2019 Apr;85(4):470–481.

24. Boddy SL, Giovannelli I, Sassani M, et al. The gut microbiome: a key player in the complexity of amyotrophic lateral sclerosis (ALS). BMC Med. 2021 Jan 20;19(1):13.

25. Nakamura R, Kurihara M, Ogawa N, et al. Investigation of the prognostic predictive value of serum lipid profiles in amyotrophic lateral sclerosis: roles of sex and hypermetabolism. Sci Rep. 2022 Feb 3;12(1):1826.

26. Neelagandan N, Gonnella G, Dang S, et al. TDP-43 enhances translation of specific mRNAs linked to neurodegenerative disease. Nucleic Acids Res. 2019 Jan 10;47(1):341–361.

27. Ma XR, Prudencio M, Koike Y, et al. TDP-43 represses cryptic exon inclusion in the FTD-ALS gene UNC13A. Nature. 2022 Mar;603(7899):124–130

28. Brown AL, Wilkins OG, Keuss MJ, et al. TDP-43 loss and ALS-risk SNPs drive mis-splicing and depletion of UNC13A. Nature. 2022 Mar;603(7899):131–137.

29. Brichta L, Shin W, Jackson-Lewis V, et al. Identification of neurodegenerative factors using translatome-regulatory network analysis. Nat Neurosci. 2015 Sep;18(9):1325–33

30. Sorenson EJ, Mandrekar J, Crum B, Stevens JC. Effect of referral bias on assessing survival in ALS. Neurology. 2007;68(8):600–2.

31. Logroscino G, Marin B, Piccininni M, et al. Referral bias in ALS epidemiological studies. PLoS One. 2018;13(4):e0195821.

