## Supplemental Tables and Figures for "The additive effect of genetic modifiers on ALS prognosis: a population-based study"

**Supplementary Table 1**. Clinical characteristics of patients carrying both *UNC13A*^C/C^ and *CAMTA1^G/G+G/T^* vs those carrying *UNC13A*^A/A+A/G^ and *CAMTA1*^T/T^

|  | ***UNC13A*^C/C^ and *CAMTA1^G/G+G/T^***  **n=68** | ***UNC13A*^A/A+A/G^ and *CAMTA1*^T/T^**  **n=1177** | **P value** |
| --- | --- | --- | --- |
| Age at onset (years, median, IQR) | 72.1 (64.1-73.5) | 67.8 (59.8-73.9) | 0.006 |
| Sex (female) | 30 (44.1%) | 526 (44.7%) | 0.92 |
| Site of onset (bulbar) | 30 (44.1%) | 368 (31.3%) | 0.032 |
| Bulbar symptoms at diagnosis | 48 (70.6%) | 639 (54.3%) | 0.029 |
| Diagnostic delay (months, median, IQR) | 8 (5.1-12) | 9 (5.1-14) | 0.51 |
| Education (median, IQR) ° | 8 (5-13) | 8 (5-11) | 0.01 |
| ALSFRS-R at diagnosis (median, IQR) | 39.5 (34.3-45) | 42 (38-45) | 0.049 |
| FVC% at diagnosis * (median, IQR) | 82 (65.5-95.5) | 90 (71-104) | 0.039 |
| BMI at diagnosis § (median, IQR) | 24 (21.5-26.4) | 24.2 (22-26.8) | 0.371 |
| ∆ALSFRS-R (points/month, median, IQR) | 0.72 (0.33-1.83) | 0.67 (0.33-1.3) | 0.177 |
| ∆Weight § (kg/month, median, IQR) | 0.65 (0.14-1.42) | 0.25 (0-0.91) | 0.0001 |
| ALS-FTD ^ | 5 (11.4%) | 140 (16.2%) | 0.394 |
| King’s stage (1/2/3/4A+4B) at diagnosis | 23/16/26/3 | 501/374/257/43 | 0.017 |
| MiToS stage (0/1/2/3/4) at diagnosis | 42/20/5/1/0 | 779/347/26/11/2 | 0.394 |
| ∆King’s | 0.23 (0.13-0.37) | 0.20 (0.10-0.349 | 0.201 |

° Available for 1238 cases (67 with *UNC13A*^C/C^ and *CAMTA1^G/G+G/T^*; 1171 with *UNC13A*^A/A+A/G^ and *CAMTA1*^T/T^)

* Available for 1161 cases (65 with *UNC13A*^C/C^ and *CAMTA1^G/G+G/T^*; 1096 with *UNC13A*^A/A+A/G^ and *CAMTA1*^T/T^)

§ Available for 1213 cases (68 with *UNC13A*^C/C^ and *CAMTA1^G/G+G/T^*; 1155 with *UNC13A*^A/A+A/G^ and *CAMTA1*^T/T^)

^ Available for 909 cases (44 with *UNC13A*^C/C^ and *CAMTA1^G/G+G/T^*; 865 with *UNC13A*^A/A+A/G^ and *CAMTA1*^T/T^)

**Supplemental Table 2**. Clinical characteristics of patients carrying both *ATXN2*^≥31^ and *CAMTA1*^G/G+G/T^ vs those carrying *ATXN2*^≤30^ and *CAMTA1*^T/T^

|  | ***ATXN2*^≥31^ and *CAMTA1^G/G+G/T^***  **n=20** | ***ATXN2*^≤30^ and *CAMTA1*^T/T^**  **n=1225** | **P value** |
| --- | --- | --- | --- |
| Age at onset (years, median, IQR) | 71.5 (64.5-76.2) | 67.8 (59.9-74.2) | 0.125 |
| Sex (female) | 6 (30%) | 550 (44.9%) | 0.18 |
| Site of onset (bulbar) | 2 (10%) | 396 (32.3%) | 0.034 |
| Bulbar symptoms at diagnosis | 10 (50%) | 677 (55.3%) | 0.88 |
| Diagnostic delay (months, median, IQR) | 5.5 (3.9-9.6) | 9 (5.1-14) | 0.017 |
| Education (median, IQR) ° | 6.5 (5-10.3) | 8 (5-11) | 0.17 |
| ALSFRS-R at diagnosis (median, IQR) | 41 (34-43.8) | 42 (37-45) | 0.99 |
| FVC% at diagnosis * (median, IQR) | 89 (79-107) | 90 (71-104) | 0.811 |
| BMI at diagnosis § (median, IQR) | 24.6 (21.8-27.3) | 24.2 (21.9-26.8) | 0.636 |
| ∆ALSFRS-R (points/month, median, IQR) | 1.08 (0.62-2.87) | 0.67 (0.33-1.34) | 0.006 |
| ∆Weight § (kg/month, median, IQR) | 0.13 (0-1.13) | 0.28 (0-0.97) | 0.493 |
| ALS-FTD ^ | 2 (16.7%) | 143 (15.9%) | 0.95 |
| King’s stage (1/2/3/4A+4B) at diagnosis | 6/7/6/1 | 518/383/277/45 | 0.72 |
| MiToS stage (0/1/2/3/4) at diagnosis | 11/8/1/0/0 | 810/359/40/12/2 | 0.82 |
| ∆King’s | 0.29 (0.20-0.63) | 0.20 (0.10-0.34) | 0.012 |

° Available for 1238 cases (20 with *ATXN2*^≥31^ and *CAMTA1^G/G+G/T^*; 1218 with *ATXN2*^≤30^ and *CAMTA1*^T/T^)

* Available for 1161 cases (18 with *ATXN2*^≥31^ and *CAMTA1^G/G+G/T^*; 1143 with *ATXN2*^≤30^ and *CAMTA1*^T/T^)

§ Available for 1213 cases (20 with *ATXN2*^≥31^ and *CAMTA1^G/G+G/T^*; 1193 with *ATXN2*^≤30^ and *CAMTA1*^T/T^)

^ Available for 909 cases (12 with *ATXN2*^≥31^ and *CAMTA1^G/G+G/T^*; 897 with *ATXN2*^≤30^ and *CAMTA1*^T/T^)

**Supplemental Table 3**. Comparison of clinical characteristics of patients carrying both *C9ORF72*^≥30^ and *CAMTA1^G/G+G/T^* vs those carrying both *C9ORF72*^≤29^ and *CAMTA1*^T/T^

|  | ***C9ORF72*^≥30^ and *CAMTA1^G/G+G/T^***  **n=38** | ***C9ORF72*^≤29^ and *CAMTA1*^T/T^**  **n=1207** | **P value** |
| --- | --- | --- | --- |
| Age at onset (years, median, IQR) | 60.4 (51.2-69.1) | 68.2 (60.3-74.4) | 0.0001 |
| Sex (female) | 16 (42.1%) | 540 (44.7%) | 0.75 |
| Site of onset (bulbar) | 14 (36.8%) | 384 (31.8%) | 0.51 |
| Bulbar symptoms at diagnosis | 17 (44.7%) | 670 (55.5%) | 0.41 |
| Diagnostic delay (months, median, IQR) | 9.5 (4.7-12) | 9 (5.1-14) | 0.35 |
| Education (median, IQR) ° | 8 (5-12) | 8 (5-11) | 0.88 |
| ALSFRS-R at diagnosis (median, IQR) | 42 (39-46) | 42 (37-45) | 0.20 |
| FVC% at diagnosis * (median, IQR) | 90.5 (71-102) | 90 (71-104) | 0.68 |
| BMI at diagnosis § (median, IQR) | 23.4 (21.6-26.6) | 24.2 (21.9-26.8) | 0.48 |
| ∆ALSFRS-R (points/month, median, IQR) | 0.67 (0.40-1.11) | 0.68 (0.33-1.35) | 0.93 |
| ∆Weight § (kg/month, median, IQR) | 0.08 (0-0.91) | 0.28 (0-0.97) | 0.41 |
| ALS-FTD ^ | 11 (34.4%) | 134 (15.3%) | 0.004 |
| King’s stage (1/2/3/4A+4B) at diagnosis | 15/20/3/0 | 509/370/280/46 | 0.012 |
| MiToS stage (0/1/2/3/4) at diagnosis | 27/10/0/1/0 | 794/357/41/11/2 | 0.60 |
| ∆King’s | 0.18 (0.15-0.34) | 0.20 (0.10-0.34) | 0.53 |

° Available for 1238 cases (36 with *C9ORF72*^≥30^ and *CAMTA1^G/G+G/T^*; 1202 with *C9ORF72*^≤29^ and *CAMTA1*^T/T^)

* Available for 1161 cases (34 with *C9ORF72*^≥30^ and *CAMTA1^G/G+G/T^*; 1127 with *C9ORF72*^≤29^ and *CAMTA1*^T/T^)

§ Available for 1213 cases (36 with *C9ORF72*^≥30^ and *CAMTA1^G/G+G/T^*; 1177 with *C9ORF72*^≤29^ and *CAMTA1*^T/T^)

^ Available for 909 cases (32 with C9ORF72≥30 and *CAMTA1^G/G+G/T^*; 877 with C9ORF72^≤29^ and CAMTA1^T/T^)

**Supplementary figures**

**Supplementary Figure 1**. Survival curves (Kaplan Meier) according to *C9orf72* GGGCC expansion. Ticks are censored patients. Median survival time: C9orf72^≥30^ 2.51 years (1.64-3.812), C9orf72^≤29^ 2.67 years (1.66-5.33), p=0.016

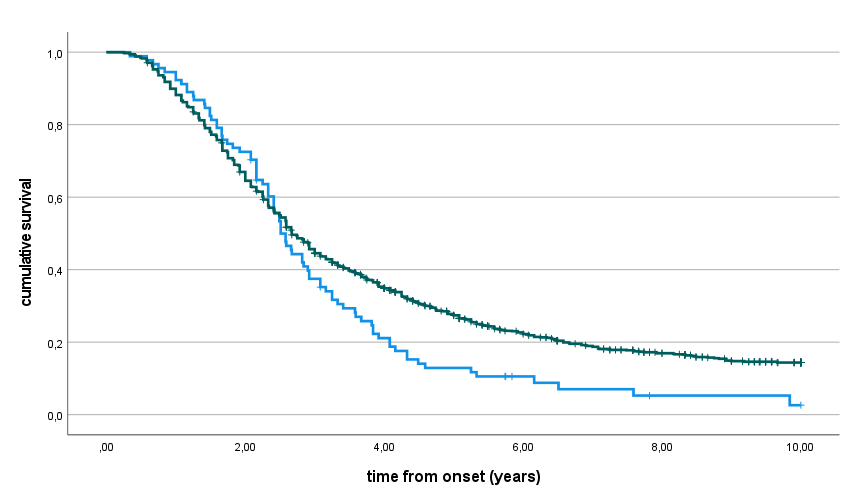

**Supplementary Figure 2**. Survival curves (Kaplan Meier) according to *ATXN2* polyQ repeats. Ticks are censored patients. Median survival time: *ATXN2*^≥31^ 1.82 years (1.080-2.330), *ATXN2*^≤30^ 2.74 years (1.67-5.26), p<0.001

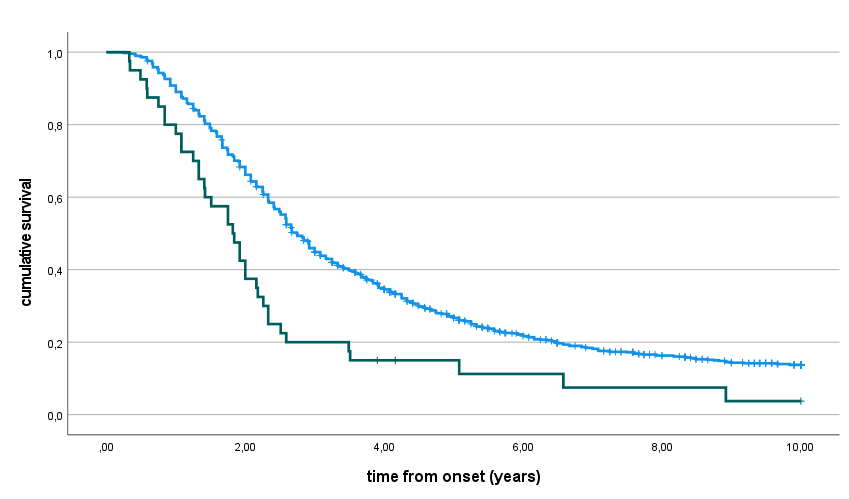

**Supplementary Figure 3**. Survival curves (Kaplan Meier) according to UNC13A rs12608932 polymorphism. Ticks are censored patients. Median survival time: *UNC13A*^C/C^ 2.25 years (1.26-4.00), *UNC13A*^A/A+A/C^ 2.82 years (1.67-5.33), p<0.001

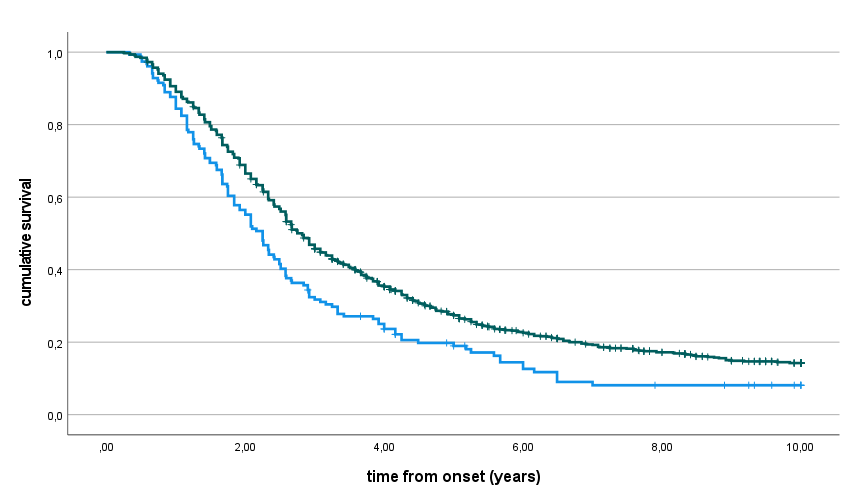

**Supplementary Figure 4**. Survival curves (Kaplan Meier) according to *CAMTA1* rs2412208 polymorphism. Ticks are censored patients. Median survival time: *CAMTA1*^G/G+G/T^ 2.58 years (1.59-5.08) vs *CAMTA1*^T/T^ 2.84 years (1.74-5.33), p=0.231

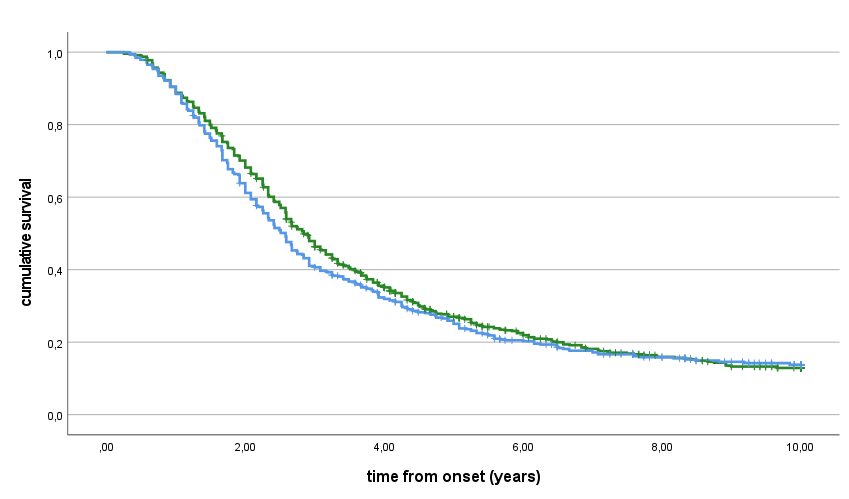

**Supplementary Figure 5**. Survival curves (Kaplan Meier) according to *SLC11A2* rs407135 polymorphism. Ticks are censored patients. Median survival time: *SLC11A2*^A/C+C/C^ 2.66 years (1.59-5.58), *SLC11A2*^A/A^ 2.74 years (1.67-4.92), p=0.665

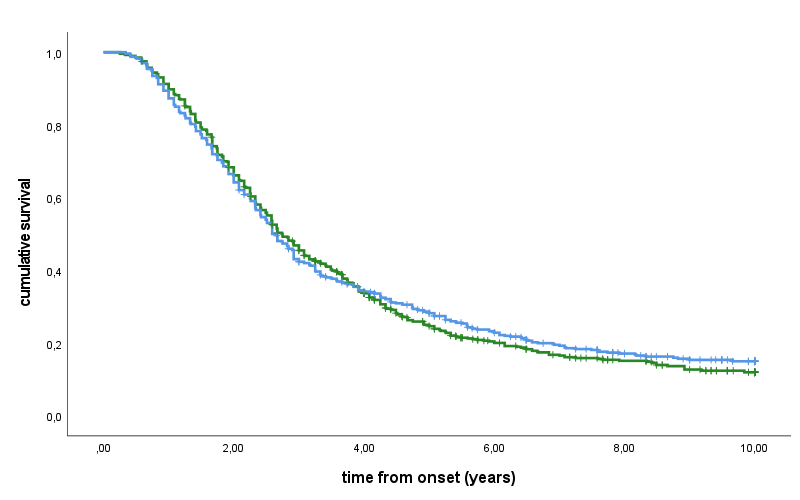

**Supplementary Figure 6**. Survival curves (Kaplan Meier) according to *ZNF512B* rs2275294 polymorphism. Ticks are censored patients. Median survival time: *ZNF512B*^C/C+C/T^ 2.66 years (1.66-5.16), *ZNF512B*^T/T^ 3.42 years (1.92-5.51), p=0.325

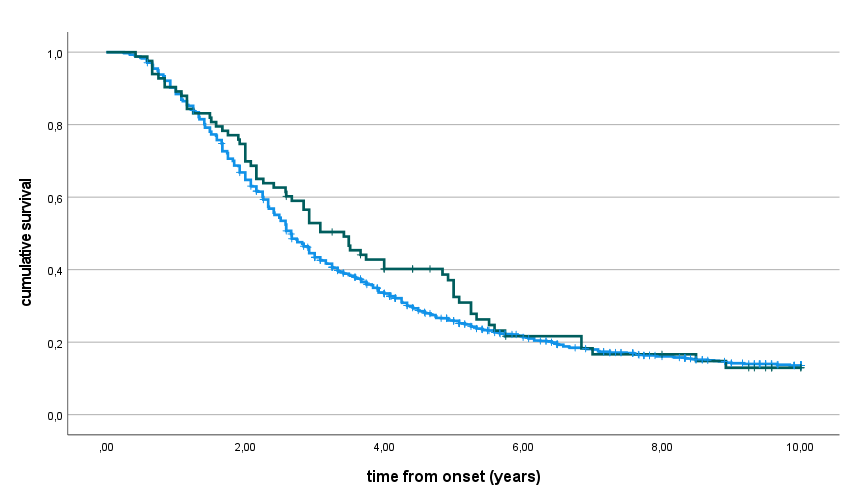
